# A novel approach to classification and segmentation of colon cancer imaging towards personalized medicine

**DOI:** 10.1101/2023.07.07.23292356

**Authors:** Keerthi Harikrishnan, Neil Botelho, Penjo Rebelo, Amit Kenkre, Amogh Tarcar

## Abstract

Recent advances in the field of pathology coupled with the rapid evolution of machine learning based techniques have revolutionized healthcare practices. Colorectal cancer accounts for one of the top 5 cancers with high incidence (126,240 in 2020) with a high mortality worldwide [1] [2]. Tissue biopsy remains to be the gold standard procedure for accurate diagnosis, treatment planning and prognosis prediction [3]. As an image based modality, pathology has attracted a lot of attention for development of AI algorithms and there has been a steady increase in the number of filings for FDA authorized use of AI algorithms in clinical practice [4]. The SemiCOL Challenge aims to develop computational pathology methods for automatic segmentation and classification of tumor and other tissue classes using H&E stained images. In this paper, we present a novel machine learning framework addressing the SemiCOL Challenge, focusing on semantic segmentation, segmentation-based whole-slide image classification, and effective use of limited annotated data. Our approach leverages deep learning techniques and incorporates data augmentation to improve the accuracy and efficiency of tumor tissue detection and classification in CRC. The proposed method achieves an average Dice score of 0.2785 for segmentation and an AUC score of 0.71 for classification across 20 whole-slide images. This framework has the potential to revolutionize the field of computational pathology, contributing to more efficient and accurate diagnostic tools for colorectal cancer.

## I. Introduction

Artificial Intelligence has transformed the field of imaging based modalities at an unprecedented speed with more than 80 FDA filings for diagnostic use(Software as a Medical Device – SaMD) in 2021 [4]. Adoption of AI based algorithms for routine pathology analysis such as biomarker quantitation and tumor grading (Prostate cancer) has led to promising development of AI applications for complex tasks for discoveries, diagnosis, prediction and prognosis of cancer. Colorectal cancer (CRC) is one of the most prevalent malignant epithelial tumors worldwide, and early and accurate diagnosis of CRC is essential for effective treatment planning and improved patient outcomes. Traditional methods of pathology are time consuming and could lead to significant intra and interob-server variability. In this study, we explore the application of machine learning to develop custom models for slide level classification as well as multi class segmentation of colon cancer images. Our methods involve building models from the scratch in lieu of pretrained models. Although, this process is computationally intensive, it allowed us to tailor these models specifically towards the task and enabled the models to learn, adapt to the unique features that were present in the different pathology images. By automating and augmenting the current workflows using AI, we can improve the speed, accuracy and consistency of the reports thereby reducing the pathologists burden. Furthermore, the use of multi-class segmentation allows the application of these models to diverse cancer types (ex: Tumor stroma, necrosis, lymph are all present across multiple cancer types) thereby broadening the clinical utility. In this paper, we describe our approach in developing, training and testing these models highlighting the transformative potential of machine learning in computational pathology and personalized medicine.

## II. Methods and Techniques

### A. Data Preparation

We found that 50%-80% of pixels provided in the manually annotated data set were unclassified. A further 3% to 18% of pixels belonged to the background class. To ensure a good balance of tissue classes and background classes careful selection of the training images would be required.

Table I shows the pixel distribution among the various classes in the annotated images provided in 3 example cases. Here class 0 denotes unannotated pixels and class 10 denotes background pixels

**TABLE I:** Pixel-wise class distribution across 3 example cases

| class | ukk_case_06 | lmu_case_08 | ukk_case_07 |
| --- | --- | --- | --- |
| 0 | 60.72% | 73.96% | 72.95% |
| 1 | 4.690% | 2.476% | 6.141% |
| 2 | 15.49% | 2.325% | 0.0% |
| 3 | 2.815% | 0.278% | 5.423% |
| 4 | 1.696% | 8.615% | 0.703% |
| 5 | 5.956% | 2.522% | 5.806% |
| 6 | 0.193% | 0.033% | 1.466% |
| 7 | 0.032% | 0.0% | 1.032% |
| 8 | 0.0% | 3.024% | 0.0% |
| 9 | 0.0% | 0.352% | 0.999% |
| 10 | 8.392% | 6.401% | 5.475% |

Each 3000×3000 image was split into 8×8 tiles and resized to 256×256 pixels. Tiles that had more than 70% of pixels belonging to the background or unannotated classes were removed.

The train and test splits were create by dividing the images on the basis of case.

### B. Training

Early attempts at training a single model to segment all 10 classes yielded models that would have high dice scores in a few classes and very poor dice scores in others. Improving the models performance on a particular class by increasing the amount of training data corresponding to that class led to a deterioration in performance of other classes. Hence 10 separate models were trained, i.e. one for each class except for the unannotated class.

A U-Net model with an EfficientNet b2 model [5] as an encoder was used for segmentation. The model had an encoder depth of 5 and a sigmoid activation at the final layer. Concurrent Spatial and Channel Squeeze & Excitation [6] was used. The model was initialized with random weights.

Given that many pixels were unannotated, updating the loss function based on these pixels may cause the model to learn incorrect features or become confused. This is because a pixel classified by the model as a tumour, but being unanotated in the ground truth would be considered an incorrect prediction even if the unannotated pixel should have been annotated as a tumour pixel. Hence a modified version of Binary Cross Entropy Loss was used to train the model such that loss was not calculated on pixels belonging to the unannotated class.

To reduce false positives only pixels with a probability higher than 0.7 were considered.

The 10 individual masks were combined using argmax, i.e for each pixel the class that had the highest probability at that pixel was assigned to it in the combined mask.

### C. Classification

Each Whole Slide Image (WSI) was systematically divided into tiles of dimensions 814 × 814 pixels, discarding any tiles that deviated from the specified shape. Subsequently, these tiles underwent resizing to 256 × 256 pixels, utilizing Bilinear Interpolation and enabling Anti-Aliasing for optimal results. This resizing is required to ensure the zoom level of WSI tiles is the same as the images the model was trained on.The resized tiles were then processed through the Tumor Segmentation model to acquire a tumour segmentation mask.

Following this, the segments were reassembled to generate a comprehensive tumor mask for the entire slide. A pixel-level threshold of 0.7 was applied to this mask, whereby any pixel with a value below 0.7 was assigned a value of 0 (with the pixel range spanning from 0 to 1).

To consider only continuous large regions of tumour, All continuous regions were extracted from the mask. The enclosing circle with minimum area was calculated for each region. Regions whose minimum enclosing circle had a radius smaller than 450 pixels were removed.

Finally, the WSI was categorized as belonging to the tumor class if the total area encompassed by the tumor within the mask exceeded 10%.

## III. Results

Table II below shows the average of the dice scores calculated on the segmentation task for 6 images from given challenge validation data.

**TABLE II:** Class wise dice score

| Class | Dice Score |
| --- | --- |
| 1 | 0.5605 |
| 2 | 0.2785 |
| 3 | 0.0896 |
| 4 | 0.5987 |
| 5 | 0.0805 |
| 6 | 0.4222 |
| 7 | 0.0411 |
| 8 | 0.0519 |
| 9 | 0.3404 |

An AUC score of 0.71 was achieved across 20 Whole Slide Images.

## IV. Conclusion

In conclusion, our study demonstrates the efficacy of denovo machine learning models at slide level classification and multi class segmentation of colon cancer images. Our approach of not using the pretrained weights, allowed us to develop new models that adapt to new histological features thereby possibly enhancing the diagnostic accuracy and reduce the inter observer variability. As we continue to refine our models to improve the accuracy, we believe our novel approach contributes to the field of computational pathology where one can envision a future for successful adoption of AI algorithms in routine clinical practice to revolutionize diagnosis, treatment and patient care.

## Data Availability

https://www.semicol.org/

## Notes

### Competing Interest Statement

The authors have declared no competing interest.

### Funding Statement

No external funding

### Author Declarations

Data was used from the SemiCOL competition as shared by the organizers. We were the Team Microscopy Mavericks and participated in this challenge and hence got access to the imaging data. https://www.semicol.org/

